# Visualising large audiological data beyond the audiogram for auditory phenotyping

**DOI:** 10.64898/2026.09.24.26363901

**Authors:** Gerard Encina-Llamas, Erik Kjærbøl, Abigail Anne Kressner

## Abstract

The pure-tone audiogram remains the cornerstone of hearing assessment, yet it captures only part of the functional consequences of cochlear damage. Most phenotyping schemes rely solely on the audiogram, which cannot detect hidden suprathreshold dysfunction when thresholds are normal. How best to represent audiological data — as discrete phenotypes or as a continuous space — remains unresolved. Using a retrospective database from the Copenhagen Hearing and Balance Centre, Rigshospitalet (Denmark; 1995–2022; 251,986 ear-session units from 81,883 adults with normal or sensorineural audiograms), we compared linear (PCA) and non-linear (UMAP) embeddings of the audiogram alone and with word recognition in quiet (WRS_max_-Q). Here we show that a linear embedding suffices for the audiogram, but a non-linear one becomes advantageous once suprathreshold measures are added. PCA compressed the audiogram onto two interpretable axes, overall level and slope, explaining 91.5% of the variance, with ears forming a continuous cloud that published reference schemes tiled into overlapping regions. For the audiogram alone, UMAP recovered the same organisation. WRS_max_-Q varied widely among ears with similar audiograms; adding it left the PCA unchanged but reorganised the UMAP. A synthetic experiment illustrated this: ears with identical audiograms but different WRS_max_-Q separated under UMAP, not PCA. Combining thresholds with WRS_max_-Q, UMAP yielded a continuous representation — a prototype *Hearing Loss Map* — where each ear’s position reflects threshold and suprathreshold measures. Representing hearing loss as a location in a continuous space rather than as discrete categories may better reflect its graded nature, with clinical relevance for individual patients.

## Introduction

For more than a century, the pure-tone audiogram has been the cornerstone of hearing assessment, and it remains the primary measurement on which most clinical decisions about hearing loss still rest. Its endurance is well earned: the audiogram is fast, reproducible, intuitive, and highly informative. Yet a substantial body of evidence now shows that it captures only part of the functional consequences of cochlear damage (see Musiek et al. 2017, for a review). Two individuals with almost identical audiograms can differ markedly in speech understanding (Grant et al. 2022), in the benefit they draw from amplification with hearing aids (Sanchez-Lopez et al. 2021), and, ultimately, in the pathology underlying their loss (Schuknecht and Gacek 1993). Indeed, sensorineural hearing loss (SNHL) is not a single, uniform condition but a heterogeneous mix of cochlear pathologies (sensory, neural, and strial/metabolic) that frequently coexist within the same ear; it is also the most common form of permanent hearing loss and a leading contributor to years lived with disability worldwide (WHO 2021). Some of these pathologies, such as inner hair cell (IHC) loss or cochlear synaptopathy, can leave the audiogram essentially untouched while presumably degrading suprathreshold function (e.g., Kujawa and Liberman 2009; Lobarinas et al. 2013). Characterising this heterogeneity in a structured and reproducible way is therefore a prerequisite for moving audiology towards precision care.

Most attempts to organise this heterogeneity have worked from the audiogram itself, along two routes. The first is mechanistic, anchoring audiometric patterns to their presumed underlying cochlear pathology. Building on the classical temporal-bone pathology of presbycusis (Schuknecht and Gacek 1993), animal models of age-related hearing loss (ARHL) have been used to define metabolic and sensory audiometric phenotypes (Dubno et al. 2013), a classification scheme later supported by demographic and longitudinal evidence (Vaden et al. 2017). More recently, this scheme has moved beyond discrete categories: the metabolic and sensory contributions can be re-expressed as continuous, additive components estimated in decibels from the audiogram, so that any given ear is described by a graded mixture rather than a single label (Vaden et al. 2022).

The second route is data-driven, letting structure emerge from the audiograms themselves. Vector quantisation of more than 28,000 audiograms yielded ten standard audiogram shapes — seven flat-to-sloping (N1–N7) and three steeply sloping (S1–S3) — now widely used as reference profiles in hearing-aid research and standardisation (Bisgaard et al. 2010); an eleventh profile, N0, a flat 0 dB HL audiogram representing essentially normal hearing, was added subsequently. At larger scale, a Gaussian mixture model (GMM) fitted to 116,400 audiograms recovered ten audiometric types and, tellingly, almost half of those records were left unclassified under conventional categorisation — exposing how poorly rigid schemes accommodate clinical variability (Parthasarathy et al. 2020). Comparable unsupervised approaches have delineated subtypes of noise-induced hearing loss (NIHL) (Wang et al. 2021). Whichever route is taken, these efforts share a defining feature: they read the pure-tone audiogram alone, and so yield what we will call *audiometric* phenotypes.

Both routes, however, read only the audiogram, which does not explicitly capture much about suprathreshold function. A separate body of work therefore extends phenotyping beyond it, and two independent efforts illustrate the move. In Denmark, the Better hEARing Rehabilitation (BEAR) project combined the audiogram with a suprathreshold test battery and, through a data-driven analysis, identified two largely independent dimensions of auditory distortion (inspired by Plomp 1978): a speech-intelligibility–related and a loudness-perception–related distortion. These define four auditory profiles (Sanchez-Lopez et al. 2018, 2020), each with a reasonably characteristic audiogram but differing in suprathreshold abilities that the audiogram does not fully capture, even among listeners who would share comparable audiometric shapes. Separately, using a different test battery and a model-based clustering approach, another group derived a larger set of interpretable profiles — thirteen in a prototypical clinical database — together with methods to harmonise and merge them across clinics and datasets (Saak et al. 2022, 2025). These studies differ in data and method, yet converge on the same conclusion: measures beyond the audiogram contain variation the audiogram cannot capture. Phenotypes of this kind are more properly described as *audiological*; we keep this distinction throughout — *audiometric* for representations built from the pure-tone audiogram, *audiological* for those that additionally incorporate suprathreshold information.

Across both lines of research, a consistent theme emerges: hearing profiles do not fall into cleanly separated clusters but populate a continuous, multidimensional space. This is already true of the audiogram itself, which records a threshold at each of many frequencies yet is well summarised by only a few dimensions. In a principal component analysis (PCA) of the hearing measures of 960 listeners, two interpretable components — overall degree of loss (PC1) and audiometric configuration, flat versus sloping (PC2) — together captured roughly three-quarters of the variance, with presbycusis phenotypes distributed continuously along them rather than in discrete classes (Allen and Eddins 2010). This favours treating phenotyping as a problem of *representation* or *segmentation* rather than of hard classification, and places dimensionality reduction at its centre. Two families of methods are available: linear techniques such as PCA, which yield interpretable axes that map onto clinically meaningful quantities such as overall level and slope (Nicolas-Puel et al. 2025), and non-linear techniques such as uniform manifold approximation and projection (UMAP), which instead preserve local neighbourhood structure and can recover geometry that a linear projection tends to flatten (McInnes et al. 2020). Which family is more appropriate need not be the same in every setting, and may depend on how much structure the data carry beyond overall level and slope. Audiogram-based profiling frameworks have been systematically compared across several large-scale cohorts and found to be broadly robust and generalisable (Xu 2026). What remains underexplored is whether these representations extend to broader audiological data beyond the audiogram, and how linear embeddings compare with non-linear ones at clinical scale — in particular, whether a non-linear embedding reveals structure that PCA cannot.

Answering this question requires large, well-curated datasets, which are now becoming available. The electronic health records (EHRs) of specialist centres already hold detailed records for tens of thousands of patients gathered over decades (e.g., Cantuaria et al. 2021). The platforms able to manage such data well have historically been costly and unevenly available, but open, purpose-built research databases are now emerging (Callejón-Leblic et al. 2024), alongside a community-wide effort to agree on common data standards and interoperable formats for audiology (Vercammen et al. 2026). As these initiatives mature, richly curated multimodal datasets, and the need for principled ways to represent and compare them, will only grow more prevalent. Exploiting them, however, depends on careful preprocessing and quality control, whose choices materially shape downstream analysis (Dillard et al. 2020).

Here, we investigate how complex audiological data at clinical scale should be represented in a low-dimensional, clinically interpretable space — and, specifically, whether the linear structure that adequately captures the pure-tone audiogram still holds once suprathreshold speech information is added. To our knowledge, this is the first systematic comparison of linear and non-linear embeddings of real-world audiological data beyond the audiogram at clinical scale. We use a retrospective database from the Copenhagen Hearing and Balance Centre (CHBC) at Rigshospitalet University Hospital (Denmark), comprising clinical diagnostic audiological examinations collected between 1995 and 2022. We first describe the composition of this dataset and the preprocessing pipeline used to render it analysis-ready, and characterise the resulting final cohort. We then evaluate PCA and UMAP as frameworks for visualising the audiometric and audiological spaces, and test our central expectation: that a linear embedding is adequate for pure-tone data (audiometric), whereas a non-linear embedding becomes advantageous once speech audiometry is included and the representation becomes *audiological* in nature and inherently more complex. To interpret these embeddings and situate our results, we benchmark them against four published reference schemes, which also shows that such schemes partition a continuum rather than isolating discrete groups; and we use a controlled synthetic example to isolate the mechanism that separates the linear from the non-linear embedding.

## Methods

### Study design and data source

This was a retrospective, single-centre, observational study of clinical audiological records from the CHBC at Rigshospitalet University Hospital (Copenhagen, Denmark) collected between January 1995 and December 2022. The study population consisted of adults with normal or sensorineural audiograms (conductive and mixed losses excluded), identified from clinical audiological records using combined audiometric and diagnostic criteria. Data were exported from the AuditBase clinical platform using Crystal Reports (SAP, Walldorf, Germany). Analyses were conducted at the ear-session level, with an ear-session defined as one ear measured at a given assessment session and treated as a separate observation. Each ear-session record comprised air-conduction (AC) pure-tone thresholds and, where performed, bone-conduction (BC) thresholds, with their masking status and levels. Records also included speech audiometry in quiet and, in a subset of ears, speech audiometry in noise, together with test metadata (test type, measurement booth, examiner and test context) and free-text clinical notes. Clinical diagnoses coded in International Classification of Diseases, 10th Revision (ICD-10) were obtained from a separate diagnostic export and linked to the audiological records by the Danish Civil Personal Registration (CPR) number and date, and assigned to ears by the laterality stated in the diagnosis (codes recorded as bilateral, or with no stated side, were applied to both ears). In addition to these clinical data, a small controlled synthetic dataset (constructed as described under Dimensionality reduction) was used to isolate, under known ground truth, the mechanism by which a non-linear embedding can separate ears that a linear one cannot.

### Audiometric and speech measures

In clinical practice at Rigshospitalet, pure-tone thresholds were obtained by standard clinical audiometry in 5 dB steps: AC (supra-aural or insert transducers) and BC, with contralateral masking when indicated. The four-frequency pure-tone average (PTA-4) is the mean AC threshold at 500, 1000, 2000 and 4000 Hz. Speech audiometry was assessed with the Danish DANTALE I monosyllabic word material (Elberling et al. 1989). In quiet, clinicians measure the speech reception threshold (SRT-Q, the level for 50% recognition) and the word-recognition (discrimination) score at its maximum across presentation levels (WRS_max_-Q), together with the level at which that maximum occurred (Level@WRS_max_-Q). In noise, clinicians measure the maximum word-recognition score in the sound field (WRS_max_-N). SRT-Q reflects audibility and is used clinically to corroborate the PTA-4, whereas WRS_max_-Q reflects suprathreshold speech processing and can differ between ears with similar audiograms. WRS_max_-N, when measured, is typically recorded in connection with hearing-aid evaluation and was available for too few ears to be used in the current study.

### Preprocessing

CPR numbers were validated (checksum and date-of-birth consistency), birth dates were reconstructed from the CPR structure, sex was derived from CPR parity, and age at examination was computed from birth and audiogram dates. Under our data-access permission, only records from adults (≥ 18 years) were available; age was nonetheless validated to confirm that no minors were included, and records were further restricted to diagnostic examinations. Examinations were grouped into measurement sessions (visits). When a session contained more than one audiogram, these were numbered chronologically by an audiogram index and combined, separately for AC and BC thresholds, into a single audiogram per session. The earliest recording was taken as the reference; any threshold missing from it was completed from later same-day recordings, where available, provided they agreed with the reference to within 10 dB at every jointly measured frequency, following the concordance criterion of Cantuaria et al. (2021). Later recordings discordant beyond this tolerance were discarded rather than used to extend the reference. This yielded one AC and, where tested, one BC audiogram per ear and examination day. Starting from 84,806 adult patients in the export, 574 were removed for an invalid or missing CPR, none for a CPR–birth-date inconsistency (27 birth dates were corrected), and 20 for a test context other than diagnostic audiometry (e.g., trial/learning or equipment testing). After merging multi-index audiograms and discarding ear-sessions without a usable audiogram, 84,063 patients remained, contributing 152,034 sessions and 295,563 ear-session units (Supplementary Figure S1).

For each ear and frequency, the threshold was taken from the masked measurement where available, and from the unmasked measurement otherwise. Missing AC and BC thresholds were then linearly interpolated across frequency and rounded to 5 dB, but only for ear-sessions in which at least three of the six octave frequencies (250, 500, 1000, 2000, 4000 and 8000 Hz) carried a measured threshold. Values beyond the measured range were held at the nearest measured threshold rather than extrapolated, and interpolated points were flagged so that observed and interpolated thresholds could be distinguished in later analyses. This preprocessing step was necessary because both the PCA and UMAP algorithms require complete observations and do not accommodate missing values.

After preprocessing and session consolidation, ears were audiometrically classified as normal, conductive, mixed, or sensorineural. Each ear was assigned an audiometric diagnosis from its air–bone gap (ABG) (AC minus BC threshold). The ABG was evaluated only at the standard octave frequencies where BC is routinely measured (250, 500, 1000, 2000 and 4000 Hz), using observed (non-interpolated) thresholds. A conductive component was considered present when the ABG reached 15 dB at two or more of these frequencies, with a stricter 20 dB required at 4000 Hz, where an isolated ABG is a common measurement artefact rather than a true conductive sign and is disregarded clinically (Cantuaria et al. 2021; Margolis et al. 2013). Each ear was then labelled *conductive* when a conductive component was present with essentially normal BC, *mixed* when a conductive component co-occurred with elevated BC (*>* 20 dB HL at two or more frequencies), *sensorineural* when no conductive component was present but AC was elevated (*>* 20 dB HL at two or more frequencies), and *normal* otherwise. Clinical ICD-10 codes were subsequently merged with the audiometric classifications. All analyses were performed at the *ear-session* level, with each observation representing a single ear from a single clinical session.

The final cohort was then defined from the 295,563 preprocessed ear-session units by applying additional diagnostic and quality-control exclusions: (i) sessions carrying any ICD-10 code from a predefined exclusion list of diagnoses not considered part of the target cohort (e.g. otosclerosis, cholesteatoma, Ménière’s disease, and otitis externa/media and related non-sensorineural conditions), removing 332 ear-session units; (ii) ears audiometrically classified as conductive or mixed (see before), removing 41,010; (iii) ear-sessions without a complete eleven-frequency AC audiogram, removing 132; and (iv) sessions with a PTA-4 ≥ 110 dB HL in either ear, removing 2103. This left 251,986 ear-session units from 81,883 unique patients (Supplementary Figure S1).

### Classification against reference schemes

To provide external reference categories for the embeddings, every ear-session was labelled against four published reference schemes: the eleven (ten original plus N0) standard audiograms of Bisgaard et al. (2010), the audiometric phenotypes of Dubno et al. (2013), the BEAR auditory profiles of Sanchez-Lopez et al. (2020), and the data-driven audiometric phenotypical clusters of Parthasarathy et al. (2020). These were originally proposed for a range of purposes — the Bisgaard set, for instance, defines standard audiograms for hearing-aid characterisation and was not intended as a patient-classification system — but we applied each here as a set of reference categories to which an ear could be assigned. An ear was assigned to a category when at least 9 of the 11 frequencies satisfied the frequency-specific criteria. When multiple categories met this criterion, assignment was based on the smallest mean absolute deviation to the category mean; non-matching ears were left unclassified. For Bisgaard we used a fixed *±*10 dB HL band around each of its eleven standard audiograms (matching the *±*10 dB coverage criterion used in their derivation); for Dubno and BEAR we used their published frequency-specific ranges; and for Parthasarathy we used strict all-frequency inclusion within its published limits, with a diagonal-covariance Mahalanobis (*χ*^2^) fall-back aligned with the GMM from which its clusters were derived. As a secondary analysis, all four schemes were additionally re-classified under one common rule (≥ 9 of 11 frequencies within *±*10 dB HL of each category’s mean, with no fallback) to evaluate them under equivalent criteria.

### Statistical analysis

To characterise the cohort, PTA-4 was related to age and mean audiometric thresholds were examined by age decade, with group means reported throughout with the standard error of the mean (SEM). To motivate the role of speech testing beyond the audiogram, each speech measure was related to PTA-4: the speech reception threshold (SRT) in quiet is expected to track the PTA-4, whereas any departure of WRS_max_-Q from PTA-4 marks word-recognition information the audiogram does not carry, and it is this residual variance that later separates the linear from the non-linear embedding. All associations were quantified by Pearson correlation (5% significance level); the two non-linear ones (PTA-4 versus age and WRS_max_-Q versus PTA-4) were additionally fitted with a two-segment (broken-stick) piecewise-linear regression with a single estimated breakpoint, capturing the accelerating loss with age and the ceiling-then-decline of word recognition, respectively. All tests were two-sided, with correlation and breakpoint estimates reported with 95% confidence intervals.

### Dimensionality reduction

#### Principal component analysis

PCA was applied to the eleven AC thresholds, mean-centred and scaled to unit variance, retaining the first two components. An initial, unconstrained PCA showed that these two components corresponded to overall hearing level and to audiometric slope (consistent with Allen and Eddins 2010; Nicolas-Puel et al. 2025; Xu 2026). Because component signs are otherwise arbitrary and can flip between runs, they were subsequently fixed to a single convention — PC1 signed to correlate positively with PTA-4 and PC2 with the high-frequency-minus-low-frequency slope (mean of 2000 – 8000 Hz minus mean of 250 – 1000 Hz) — so that severity increases along Dimension 1 and slope along Dimension 2 in every analysis.

#### Uniform Manifold Approximation and Projection

UMAP was fitted on manually *z*-scored features (means and standard deviations stored so new points can be projected consistently), with 2 components, 100 neighbours, minimum distance of 0.80, 1000 epochs, Euclidean metric, PCA initialisation, and learning rate of 1.5. UMAP dimensions were oriented to the PCA convention by sign-flipping against the same PTA-4 and slope anchors, solely to aid interpretation and to allow direct comparison with PCA. Two feature sets were used: *tone-only* (the eleven AC thresholds) and *tone+speech* (the eleven AC thresholds plus WRS_max_-Q), each restricted to complete cases (251,986 and 213,237 ear-session units, respectively). Reference-scheme mean audiograms were projected into the fitted PCA and UMAP spaces for overlay.

#### PCA versus UMAP

To compare the two methods, both were fitted on the identical tone-only and tone+speech complete-case subsets, giving four two-dimensional embeddings shown with common axis limits within each method and coloured by an example reference scheme, the Bisgaard standard audiograms.

#### Synthetic data analysis

To isolate, under known ground truth, the mechanism by which a non-linear embedding can separate ears that a linear one cannot, a controlled synthetic dataset was generated from the Bisgaard reference profiles: for each standard audiogram (N0–N7, S1–S3) an equal number of 1000 synthetic audiograms was drawn uniformly within its *±*10 dB band. Each ear was then assigned a WRS_max_-Q score from a profile-specific normal distribution. These distributions were a simplified approximation of the empirical per-profile WRS_max_-Q means and spreads in the cohort, collapsed into severity-graded bands (mean, SD in %): (95, 5) for N0–N2 and S1, (75, 10) for N3 and S2, (55, 15) for N5 and S3, (40, 20) for N6, and (25, 20) for N7, with scores clipped to 0 – 100 %. Profile N4 was the deliberate exception: despite having identical audiograms, half of its ears were drawn from a good scores distribution (75, 5) and half from a poor one (25, 5), simulating a good-ear/bad-ear split beyond the audiogram. PCA and UMAP were then applied exactly to the synthetic datasets as for the clinical data, tone-only and tone+speech.

#### Software and reproducibility

Analyses were performed in R (version 4.6.0). PCA used stats::prcomp, UMAP used uwot, piecewise regression used segmented, and figures used ggplot2 with patchwork. Fitted models with their scaling parameters were cached so that embeddings and projections are reproducible. The pipeline was parallelised and run on a high-performance computing cluster (Computerome).

#### Approvals and data governance

Under Danish law, retrospective research on existing clinical data does not require informed patient consent or approval from a research-ethics committee. The study was authorised by the Center for Regional Udvikling (Capital Region of Denmark) under Pactius project P-2022-629 and conducted in accordance with the Declaration of Helsinki and the General Data Protection Regulation (GDPR). Records were linked through the Danish CPR number. All data access and analysis took place within the secure, GDPR-compliant DTU Computerome high-performance computing facility, under a data-access agreement between Rigshospitalet and DTU Computerome and restricted to two named researchers (G.E.-L. and A.A.K.). Only aggregated, de-identified outputs left this environment.

## Results

### Dataset composition and demographics

The database drawn from the CHBC at Rigshospitalet University Hospital comprised diagnostic audiological assessments from 84,806 unique adult patients seen between January 1995 and December 2022 (Figure 1). After curation and quality-control preprocessing (see Methods; Supplementary Figure S1), 84,063 patients remained, contributing 152,034 sessions (each a clinical visit that included audiological testing) and 295,563 ear-session units.

**Figure 1.**
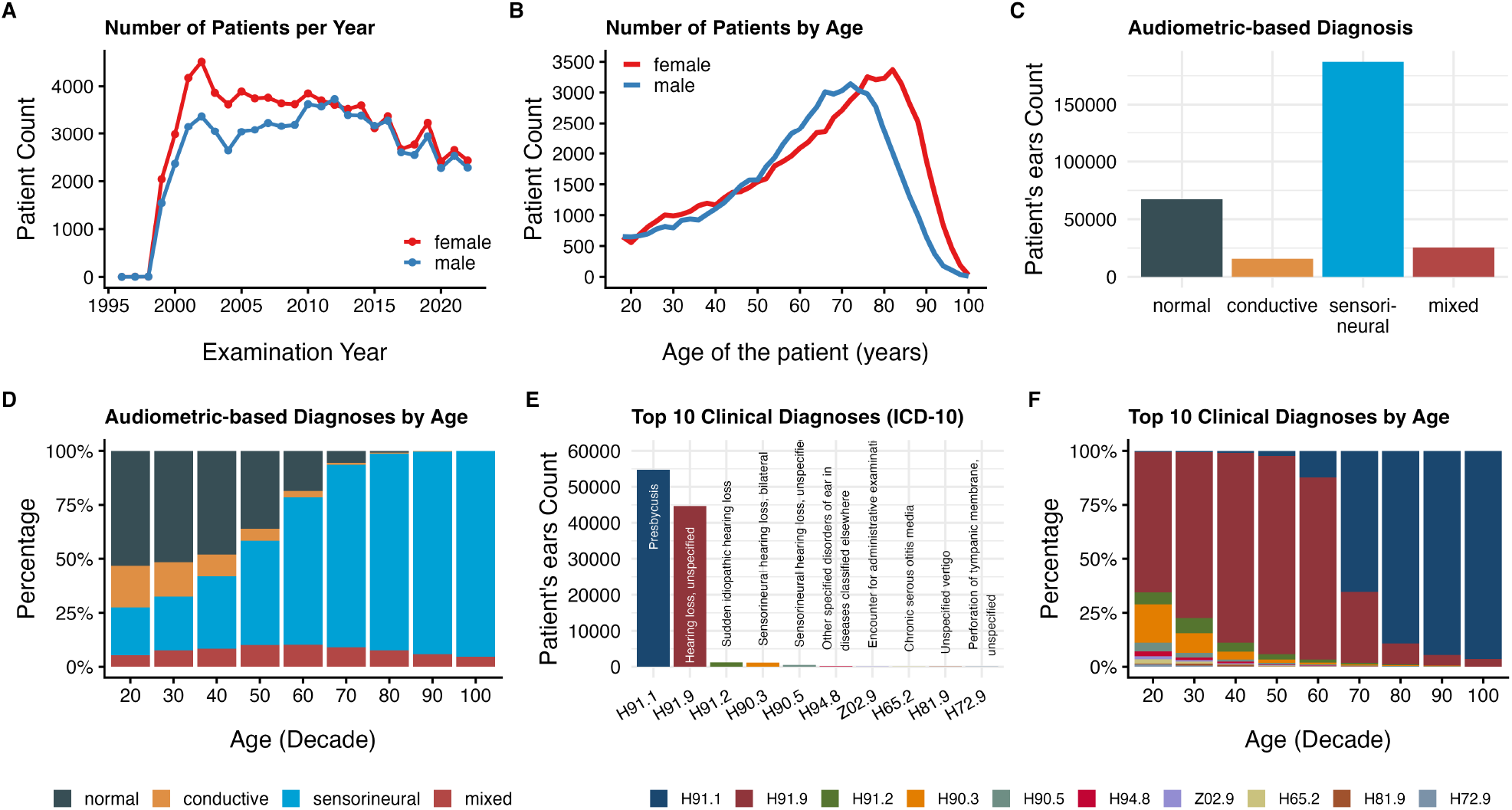
General description of the data (1995–2022). (A) Number of patients examined per year, by sex (female in red, male in blue). (B) Number of patients by age, by sex (female in red, male in blue). (C) Distribution of audiometrically derived diagnosis categories (normal, conductive, sensorineural, mixed) across all ear-session units. (D) Relative frequency of audiometric diagnoses by age decade. (E) Counts of the ten most frequent ICD-10 clinical diagnosis codes. (F) Relative frequency of the ten most frequent ICD-10 codes by age decade.

Recording on the AuditBase platform began in 1995, but annual examination numbers rose steeply only from 1998 and peaked in 2002 before declining gradually; women slightly outnumbered men throughout, most visibly between 2000 and 2010 (Figure 1A). The number of patients rose monotonically with age to a peak at 73 years in men and 82 years in women, then fell sharply (Figure 1B). This age distribution is consistent with a specialist ear, nose and throat (ENT) population dominated by ARHL, although a substantial proportion were younger, with 25.7% below 50 years. Diagnoses were predominantly sensorineural (63.3% of ear-session units), with smaller fractions normal (22.8%), conductive (5.3%), or mixed (8.6%) (Figure 1C). The proportion of SNHL rose steeply with age, from about 22.2% in the third decade of life (20s) to most ears (91.2%) by the ninth decade of life (80s) (Figure 1D). Clinical ICD-10 codes, which record the clinician’s per-visit diagnosis and are lateralised only where a side was stated, were dominated by presbycusis (H91.1) and unspecified hearing loss (H91.9): together these accounted for 96.3% of ear-session units with a clinical diagnosis code (99,338 of 103,148 coded ear-session units; the remainder carried no code), followed by sudden idiopathic hearing loss (H91.2) and other sensorineural codes (Figure 1E). The coded diagnoses shifted markedly towards presbycusis with age (Figure 1F). That the two most frequent codes, presbycusis and unspecified hearing loss, are both coarse, non-specific labels in an ageing clinical population underscores the limited specificity of routine clinical diagnosis and motivates the data-driven audiological characterisation that follows.

### Cohort description

The cohort was defined by excluding sessions with an ICD-10 code outside the target cohort definition (e.g., otitis media, cholesteatoma, Ménière’s disease), ears audiometrically classified as conductive or mixed, ears without a complete AC audiogram, and sessions with a PTA-4 ≥110 dB HL in either ear. This left 81,883 patients and 251,986 ear-session units, of which 26.7% were classified via their audiogram thresholds as having normal hearing and 73.3% as having sensorineural hearing loss.

Mean AC audiograms by age decade followed the canonical pattern of presbycusis: near-normal low-frequency thresholds in the youngest decades and progressive high-frequency loss that deepened with age, reaching roughly 65–80 dB HL at 4000 – 8000 Hz in the oldest groups (Figure 2A). Among the speech measures (Figure 2B, left and right ears plotted separately), SRT-Q and WRS_max_-Q were the most widely available, each recorded in about 106,000 ear-session units per side (from about 73,000 patients); Level@WRS_max_-Q in roughly 70,000 per side (approximately 52,000 patients); and WRS_max_-N in only a few hundred (450 – 730 ear-session units per side from about 1100 patients). SRT-Q clustered at low-to-moderate values, mirroring the spread of mean pure-tone thresholds, whereas WRS_max_-Q concentrated near the 100% ceiling with a tail towards poorer scores, as expected for a cohort spanning normal to impaired ears.

**Figure 2.**
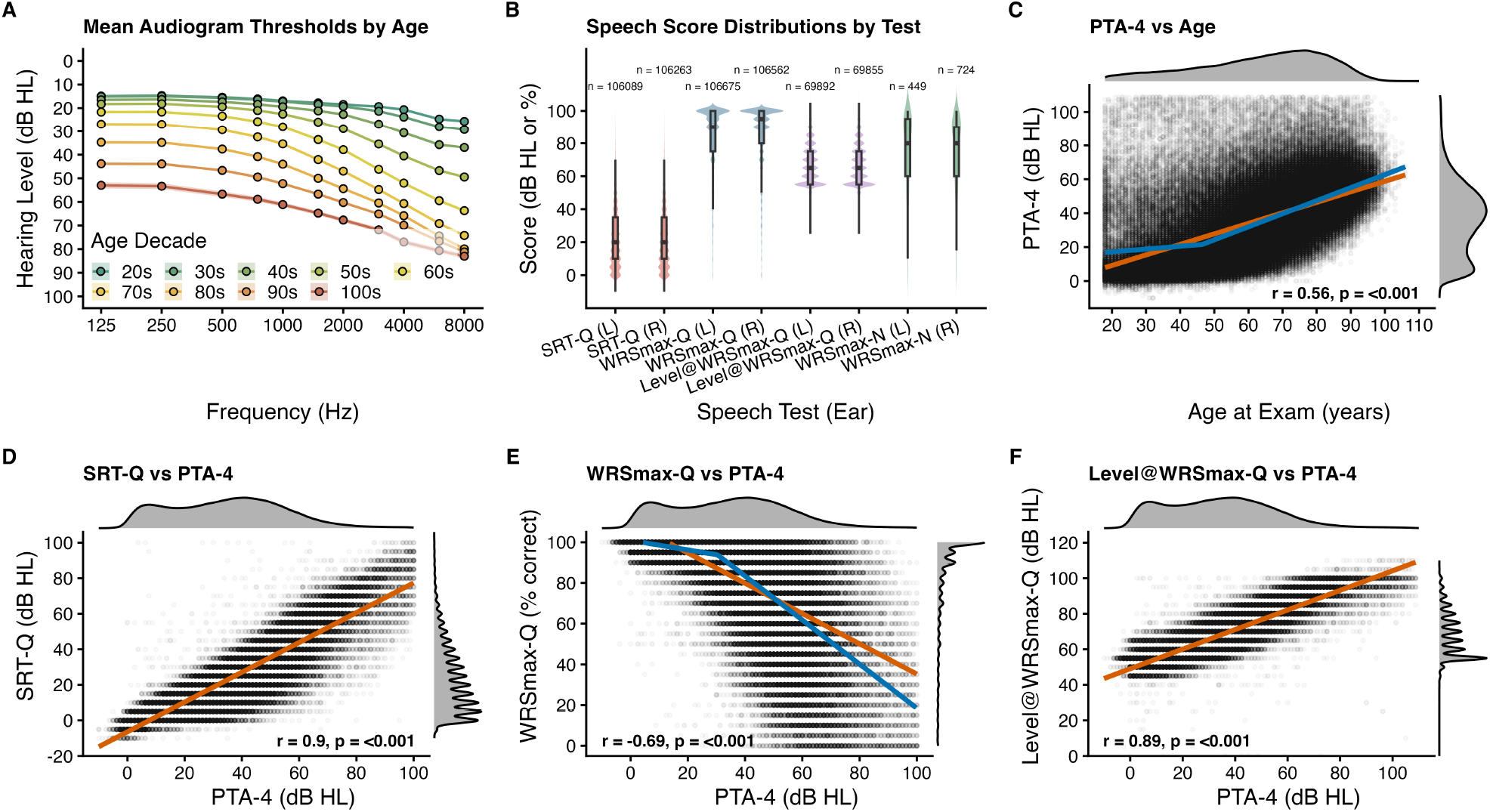
Audiometric and speech characteristics of the final cohort. (A) Mean AC audiograms by age decade (*±*SEM), coloured from younger (green) to older (red). (B) Distributions of the speech measures (SRT-Q, WRS_max_-Q, Level@WRS_max_-Q, and WRS_max_-N) for left and right ears, as violins with embedded boxplots; the number of ear-session units per ear side is annotated for each test. (C) PTA-4 versus Age at examination. (D) SRT-Q versus PTA-4. (E) WRS_max_-Q versus PTA-4. (F) Level@WRS_max_-Q versus PTA-4. Scatter panels (C–F) show point-density scatter with marginal distributions and a linear (Pearson) fit as an orange line, with *r* annotated (*p <* 0.001 throughout); panels (C) and (E) additionally show a two-segment piecewise-linear fit as a blue line.

The tone and speech audiometry measures were related but not fully redundant, which matters for the analyses that follow. Overall, PTA-4 rose with age (*r* = 0.56, *p <* 0.001; Figure 2C), but this association was strongly age-dependent. A two-segment piecewise-linear fit (blue line) placed a breakpoint at 46.5 years (95% CI 46 – 47 years): below it, thresholds were essentially flat with age (0.17 dB HL per year), rising roughly 4.5-fold faster above it (0.77 dB HL per year). Indeed, before the mid-forties the age association was effectively absent; the modest overall correlation thus reflects the steep threshold increase from mid-life onward, consistent with the onset of presbycusis. SRT-Q tracked PTA-4 very closely (*r* = 0.90; Figure 2D), as did the level employed to assess word recognition in quiet (*r* = 0.89; Figure 2F). Maximum word recognition itself declined only moderately with severity, with large residual scatter (*r* = *−*0.69; Figure 2E): a two-segment fit (blue line) showed it holding near ceiling up to a PTA-4 of 30.4 dB HL (95% CI 30.1 – 30.7 dB HL) and then falling roughly five-fold faster (−0.23 versus −1.08% per dB); beyond about 40 dB HL, two ears with the same PTA-4 could differ from near-ceiling to markedly reduced word recognition. Such variation in suprathreshold word recognition among ears with similar audiograms reflects information beyond the audiogram itself and therefore cannot be captured by a purely audiometric representation.

### Visualising audiometric data with a linear embedding (PCA)

Before exploring this suprathreshold information further, we first ask how the audiogram alone is organised. Every ear-session unit was labelled under all four published reference schemes to provide external reference categories for the embeddings that follow (Supplementary Table S1).

Coverage of the cohort was uneven: the fraction of ear-session units left unclassified was 38.8% for Bisgaard, 37.3% for Dubno, 39.8% for BEAR and 11.7% for Parthasarathy. Because the four schemes were constructed differently and each was applied under its own published criteria, these percentages are not strictly comparable. Re-classifying all four under a single common rule (at least nine of eleven frequencies within *±*10 dB HL of each category’s mean) left far more ears unclassified — 67.8% for Dubno, 71.0% for BEAR and 53.0% for Parthasarathy, while Bisgaard, already using this tolerance, was unchanged (38.8%). Further analyses are therefore based on the original definition of each scheme, which classifies the largest share of ears and gives the most complete phenotype overlays. How the four schemes relate to one another, and to the underlying variation in the audiograms, becomes clear only once every ear-session unit is placed in a common low-dimensional space for visualisation, which is the focus of the subsequent analyses.

PCA of the 11 AC thresholds compressed the audiogram onto two dominant axes: the first captured 77.7% of the variance and the second 13.8%, together 91.5% of the total (Figure 3), representing most of the data variance (Supplementary Figure S2B). Dimension 1 (abscissa) indexes overall hearing level, increasing eastward, and Dimension 2 (ordinate) audiometric slope. This interpretation is borne out by the frequency loadings, showing that all audiometric frequencies contribute to the first component while high frequencies contribute more to positive values in the second component and low audiometric frequencies to negative PC2 values (Supplementary Figure S2A). More directly, projecting synthetic audiograms into the space shows that flat audiograms of increasing level (−10 to 100 dB HL) move along Dimension 1, whereas sloping (high-frequency loss) and rising (low-frequency loss) audiograms are displaced towards positive (north) and negative (south) values of Dimension 2, respectively (Supplementary Figure S2C). Ears also order by age along Dimension 1, as expected for a severity axis (Supplementary Figure S2D). Individual ear-session units form a single continuous, island-like cloud rather than distinct clusters.

**Figure 3.**
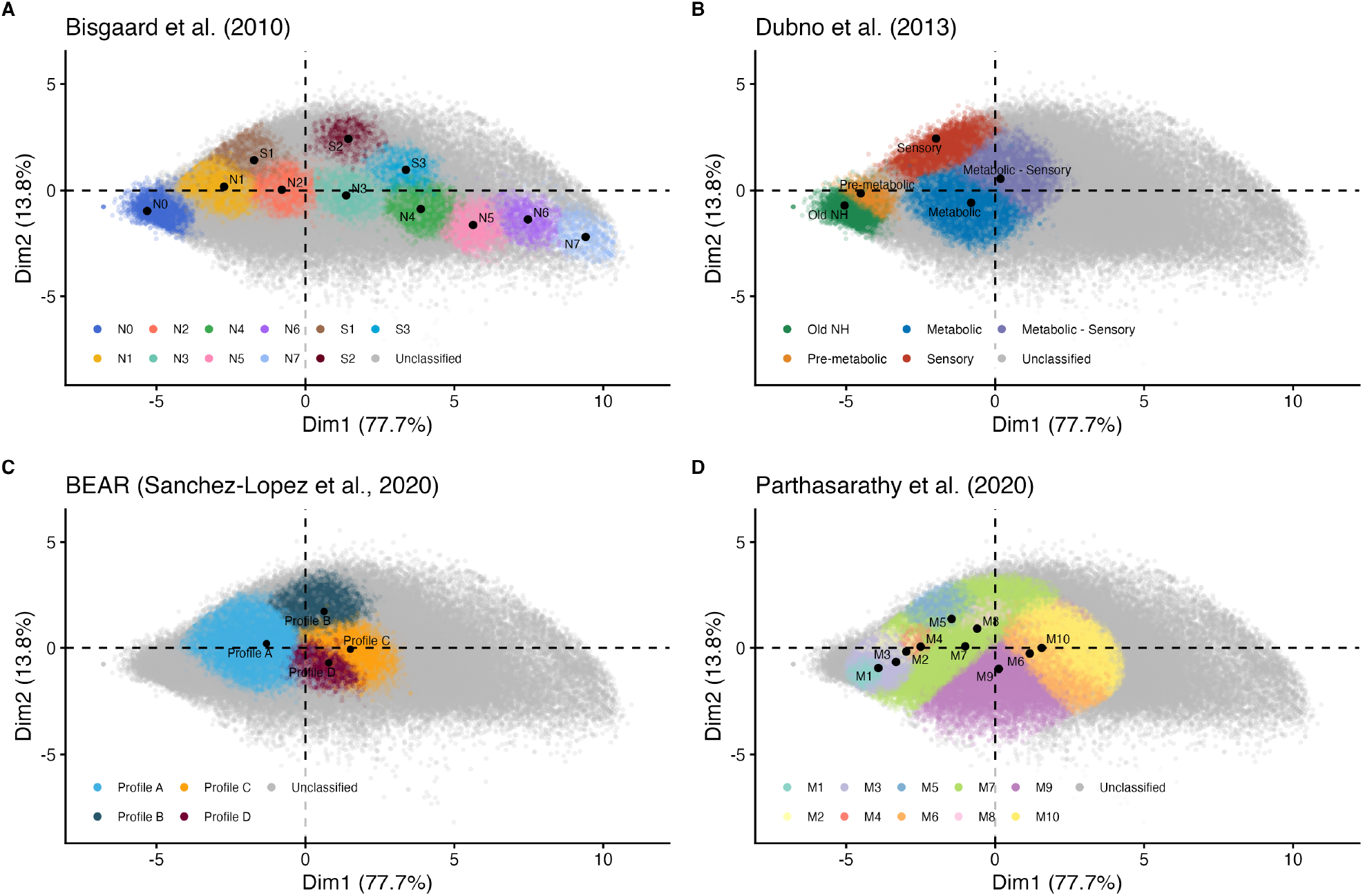
PCA of the final cohort using the eleven AC thresholds (125 – 8000 Hz), with published reference schemes overlaid. Every panel shows the same score plot (Dimension 1, 77.7% of variance; Dimension 2, 13.8%), coloured by one scheme with class centroids marked in black and unclassified ears in grey: (A) Bisgaard et al. (2010); (B) Dubno et al. (2013); (C) BEAR (Sanchez-Lopez et al. 2020); (D) Parthasarathy et al. (2020). Dimension 1 increases with PTA-4 (greater hearing loss to the east); Dimension 2 with the high-frequency-minus-low-frequency slope (sloping audiogram to the north; rising audiogram to the south).

Overlaying the classifications from each reference scheme onto the PCA representation shows the same continuous space segmented in different ways, with the ear-session units that each scheme leaves unclassified shown in grey (Figure 3A–D). The Bisgaard profiles run along Dimension 1 in order of severity, from N0 in the west to N7 in the east, with the steep S-series displaced to the north (Figure 3A). The Dubno phenotypes follow the same geometry: old-normal and pre-metabolic ears in the west, the sensory phenotype to the north (high-frequency losses), the metabolic phenotype in the centre (flatter losses), and the metabolic-sensory phenotype between the two (Figure 3B). The BEAR Profile A spans the low-to-moderate range in the west, with Profiles B, C and D clustered near the centre (Figure 3C). The ten Parthasarathy clusters span the space from near-normal (M1–M3) in the west to severe (M10) in the east, with clusters differing in area and overlapping to varying degrees (Figure 3D). In every case, the class centroids are distinct, yet the surrounding coloured point clouds overlap continuously throughout the PCA space.

### Visualising audiological data with a non-linear embedding (UMAP)

The PCA results suggest that audiograms occupy a largely continuous low-dimensional space. To examine whether a non-linear embedding would reveal additional structure within this space, UMAP (McInnes et al. 2020) was applied to the same data. For audiometric thresholds alone, the two methods yielded broadly similar results: UMAP reproduced the overall organisation seen in the PCA representation, albeit with some distortion of the embedding geometry and the Bisgaard profile boundaries, while preserving Dimension 1 as an axis of overall severity and Dimension 2 as an axis of audiometric slope (Figure 4A,C). Likewise, the four reference schemes occupy the same relative positions as in the PCA representation (Figure 3 vs Supplementary Figure S3). Thus, for audiometric data alone, the non-linear embedding revealed no major organisational structure beyond that already captured by the linear projection.

**Figure 4.**
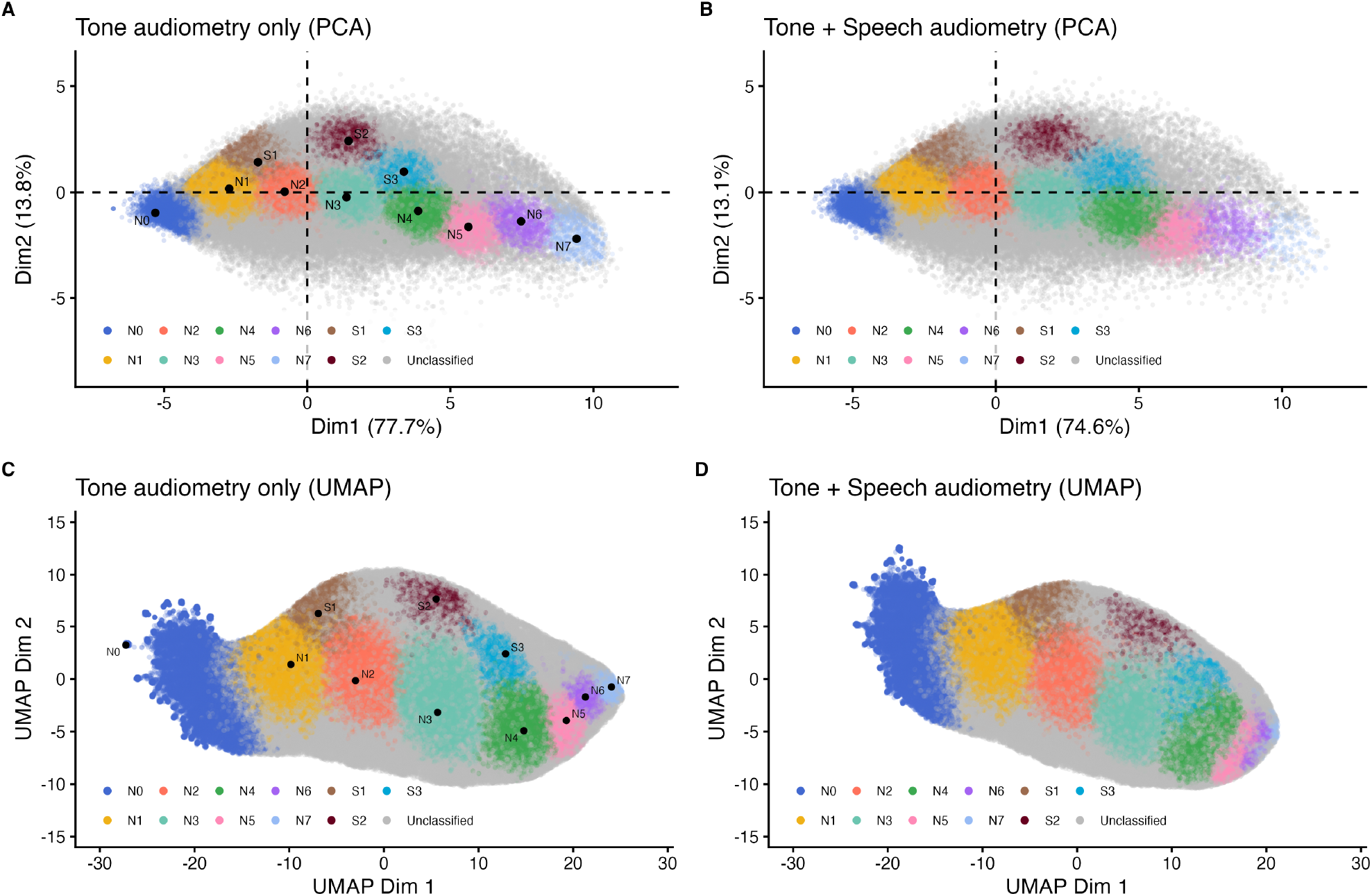
Comparison of PCA and UMAP for tone-only versus tone+speech features, with Bisgaard colouring. (A) PCA, tone-only. (B) PCA, tone+speech. (C) UMAP, tone-only. (D) UMAP, tone+speech. Axis limits are aligned within each method to show the effect of adding the maximum word-recognition score in quiet (WRS_max_-Q).

The agreement between PCA and UMAP held only for audiometric data alone. Once suprathreshold word recognition was added, the two embeddings behaved differently. In the subset of ear-session units with complete pure-tone threshold and speech audiometry data (213,237 of 251,986), adding WRS_max_-Q left the PCA essentially untouched: the overall cloud and the position and size of the Bisgaard profiles were unchanged, and the variance split barely moved (Dimension 1 from 77.7% to 74.9%; Figure 4A,B). The only visible change was a lower point density along the high-severity edge (eastern part of the island), where fewer impaired ears carry a speech score. The linear projection therefore captures little of the added information. In contrast, UMAP was sensitive to the added speech information: the overall embedding became slightly rotated and compressed along Dimension 2. Although the profiles kept their overall arrangement, some of them spread over a larger area (e.g., S3 and N4) and others contracted (N5–N7), and several overlapped more than in the tone-only audiometry embedding (Figure 4C,D). This change was concentrated in the central and eastern part of the map, over the more-impaired profiles (N3–N7, S2, and S3), where word recognition varies most widely across ears with the same audiogram (Figure 2E).

To better understand how speech information influences the embedding, we next turned to a controlled synthetic example with known ground truth (Figure 5). Synthetic audiograms were drawn from the Bisgaard reference scheme (each profile’s mean and range), and every synthetic ear was assigned a word-recognition score from a realistic, profile-specific distribution (inspired by the real data), with the exception of profile N4 in which half of the ears received good WRS_max_-Q scores and the other half poor scores despite identical audiograms (Supplementary Figure S4). As with the clinical data, PCA represented the synthetic data in essentially the same way with and without the speech scores, aside from a very subtle widening of the region occupied by N4 (Figure 5B). UMAP, by contrast, placed the synthetic profiles differently once the speech scores were added (Figure 5D): the area and shape occupied by each profile reflected its WRS_max_-Q distribution, with narrower distributions giving more compact clusters and wider distributions more elongated, larger ones, while the overall placement stayed consistent (severity increasing along Dimension 1, N0 to N7; sloping audiograms at higher values of Dimension 2, S1–S3). Most notably, profile N4, whose scores were split into a good and a poor distribution while maintaining an identical audiogram distribution, appeared as two separate green clusters (Figure 5D): one near S3 and N5 (good scores) and the other further away, near N6 and N7 (poor scores). Together, these observations suggest that a linear projection is adequate for representing pure-tone audiograms, whereas a non-linear embedding becomes beneficial once suprathreshold information is incorporated. Building on this, we propose representing hearing loss as a location in a continuous, interpretable space rather than a discrete category: the starting point for a *Hearing Loss Map*.

**Figure 5.**
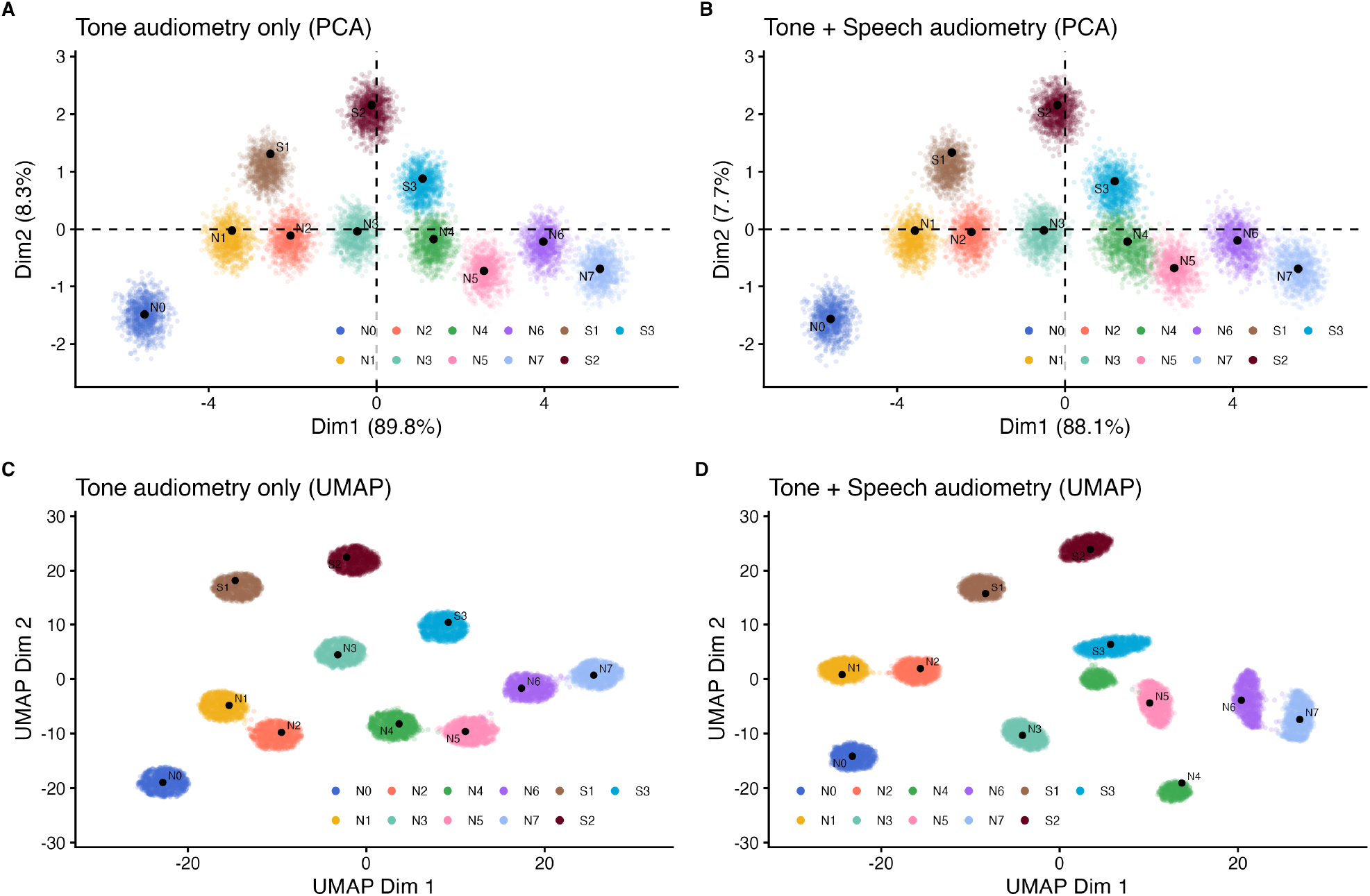
Synthetic illustration of why a non-linear embedding becomes necessary once word recognition varies independently of the audiogram. Audiograms were generated from the Bisgaard reference profiles and each ear assigned a realistic, profile-specific WRS_max_ score, except profile N4, split into good- and poor-scoring halves at an identical audiogram (Supplementary Figure S4). (A) PCA and (C) UMAP of the tone audiograms only; (B) PCA and (D) UMAP after adding WRS_max_. PCA keeps the bimodal N4 profile in a single region, whereas UMAP separates its two halves by word recognition.

### Towards a Hearing Loss Map with a UMAP-based non-linear embedding

Having established that a non-linear embedding can incorporate suprathreshold information not captured by the audiogram alone, we next examine the resulting representation. Combining the audiometric thresholds with word recognition scores (WRS_max_-Q), UMAP yields a single two-dimensional representation of the audiological space (Figure 6). Its overall organisation mirrors the audiometric embeddings: severity runs along Dimension 1, from better (west) to poorer (east) thresholds, and audiometric slope along Dimension 2, with sloping high-frequency losses to the north and rising profiles to the south. Every ear occupies a single location, and the four published reference schemes again tile the space continuously. Unlike the audiometric embeddings, however, this representation is shaped by suprathreshold performance as well as by the audiogram. We propose this representation as a prototype *Hearing Loss Map*, in which hearing loss, reflecting both threshold and suprathreshold information, is a location within a continuous space rather than a member of a discrete category. This representation has the potential to reveal audiological differences that the pure-tone audiogram, and the categories built on it, currently leave hidden.

**Figure 6.**
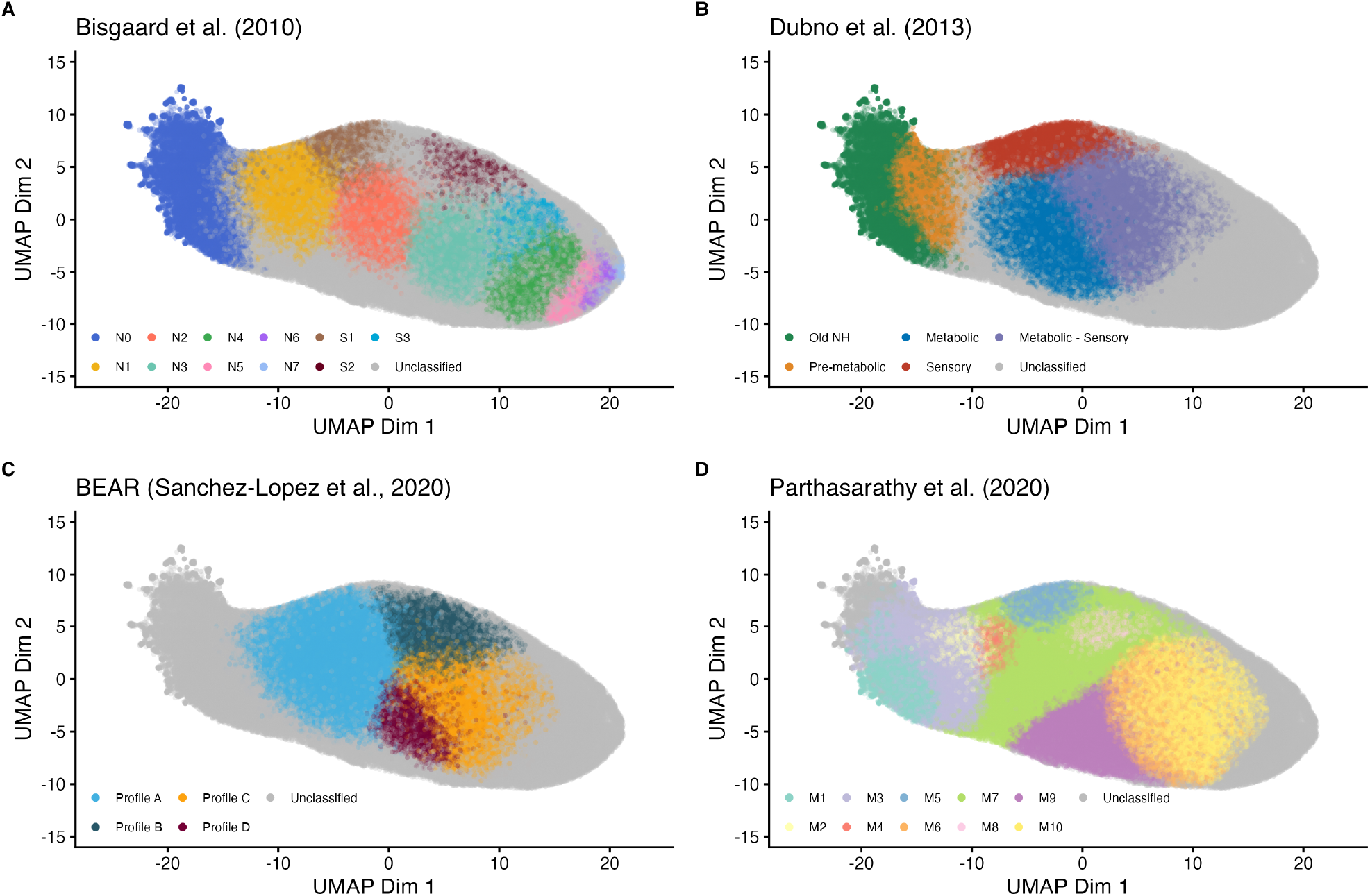
UMAP embedding of the final cohort using the eleven air-conduction thresholds together with word recognition (WRS_max_-Q), with published reference schemes overlaid. Panels and colouring mirror Figure 3: (A) Bisgaard; (B) Dubno; (C) BEAR; (D) Parthasarathy. Dimension 1 is signed to increase with PTA-4; Dimension 2 with the HF–LF slope.

## Discussion

The pure-tone audiogram carries less independent information than its individual frequencies suggest. In a clinical cohort of 251,986 ear-session units, two dimensions, overall level and slope, accounted for 91.5% of the total variance, and the ears formed a single continuous cloud rather than separating into natural groups. A linear embedding (PCA) captured most of this structure; a non-linear one (UMAP) added little. The picture changed only once the representation ceased to be purely audiometric. Word recognition in quiet (WRS_max_-Q) was related to the audiogram but not fully determined by it. That residual variance, orthogonal to level and slope, was invisible to the linear projection yet reshaped the non-linear one. The key distinction is therefore not between linear and non-linear methods but between kinds of data: a linear embedding suffices while the representation is audiometric, and a non-linear one becomes advantageous once it turns audiological.

### Building an analysis-ready cohort from routine clinical data

Results of this kind are only as reliable as the records behind them. Routine audiological data are shaped by why each test was ordered, by changes in equipment and staff over the years, and by the particular habits of a single clinic, so turning them into an analysable cohort required a long series of decisions (Dillard et al. 2020), much as the Hearing Examinations in Southern Denmark database did (Cantuaria et al. 2021). Preprocessing and the exclusion of conductive and mixed ears produced the final cohort of normal-hearing and sensorineural ears; because none of these steps is neutral, we report the attrition at every stage (Supplementary Figure S1). Reassuringly, a much larger clinical cohort, built independently, reached its final sample through almost the same choices (Clifford et al. 2026). Our cohort also reflects its origin in a specialist ENT service in Denmark: examinations declined after 2002, probably as some patients moved to private clinics, and men reached their peak numbers earlier than women (near 73 versus 82 years). This sex difference is consistent with known differences in noise exposure and care-seeking, also seen in large normative samples (Wasano et al. 2021). The onset of hearing loss with age also matched expectation: PTA-4 rose sharply only after about 46.5 years (Figure 2C), consistent with the accelerating high-frequency loss of presbycusis (Wasano et al. 2021; Clifford et al. 2026).

### The audiogram is intrinsically low-dimensional

The low dimensionality of the audiogram is not specific to our sample; it is a property of the measurement itself. Mean audiograms in our cohort followed the typical pattern of ARHL: near-normal at low frequencies in younger people, and progressively worse at high frequencies with age. This is the same pattern reported in normative samples (Wasano et al. 2021), in a very large clinical cohort (Clifford et al. 2026), and in the populations in which audiometric phenotypes were first defined (Dubno et al. 2013; Allen and Eddins 2010; Grant et al. 2022). Because thresholds at neighbouring frequencies are strongly correlated, almost all of this variation resolves into two directions: overall level, and the balance between low and high frequencies (the slope). Other studies reach the same two axes by different means. Allen and Eddins (2010) recovered them by applying PCA to audiometric and otoacoustic measures and, like us, found a continuum without natural groups. Additionally, Vaden et al. (2022) described continuous metabolic and sensory patterns of ARHL occupying a similar conceptual space. The same two directions recur whenever audiograms are reduced via a linear embedding, even for unrelated purposes such as predicting balance dysfunction (Nicolas-Puel et al. 2025), and remain stable when compared across datasets (Xu 2026). Our PCA, with severity along Dimension 1 and slope along Dimension 2, therefore recovers the geometry of the audiogram rather than imposing one.

### Word recognition carries information the audiogram does not

Speech testing tells a different, and more informative, story. The SRT followed the PTA-4 almost exactly (*r* = 0.90), confirming that in quiet it reflects little more than audibility (Figure 2D). WRS_max_-Q behaved differently: it stayed near ceiling until the PTA-4 reached about 30 dB HL and then declined (Figure 2E), but with such wide scatter that beyond 40 dB HL two ears with the same PTA-4 could range from near-perfect to markedly reduced word recognition (*r* = *−*0.69). Dubno et al. (1995) formalised exactly this scatter as confidence limits on the maximum score expected for a given loss, and the ears that fall below those limits have since acquired a mechanistic interpretation: word-recognition deficits that cannot be explained by audibility are associated with cochlear neural degeneration, seen both in clinical databases comparable to ours (Grant et al. 2022; Clifford et al. 2026) and in temporal-bone studies relating poor word scores to primary neural loss (Wu et al. 2021). At least part of this scatter is therefore unlikely to be mere measurement noise: it is consistent with the suprathreshold dysfunction that an apparently normal audiogram can hide (e.g., Kujawa and Liberman 2009; Lobarinas et al. 2013), and it is exactly the information that a purely audiometric representation, however carefully built, cannot capture.

### Linear suffices for the audiogram, non-linear for the audiological space

These two observations, a low-dimensional audiogram and a word-recognition score that it cannot fully predict, explain why the choice of embedding mattered only in some cases. For the audiogram alone, the linear and non-linear embeddings were essentially equivalent: PCA and UMAP arranged the ears similarly and placed the four reference schemes in the same relative positions (Figures 3, 4A,C, and Supplementary Figure S3). The greater complexity of a non-linear method conferred no advantage. Adding word recognition (WRS_max_-Q) broke this equivalence. Because its variance was largely orthogonal to the two dominant PCA axes, level and slope, it loaded on a minor component and left the PCA almost unchanged (Figure 4A,B). UMAP, which preserves neighbourhood structure rather than ordering directions by variance, reorganised in response, mainly over the more impaired profiles (N3–N7, S2, S3), where word recognition varies most among ears with the same audiogram (Figure 4C,D). A synthetic experiment isolated the mechanism (Figure 5). With the audiogram held fixed and only profile N4 split by word recognition (Supplementary Figure S4), PCA kept the two halves together while UMAP separated them. This confirms that a non-linear embedding can preserve information that a linear one discards. We are cautious, however, not to over-interpret the clinical data: UMAP did not resolve ears with the same audiogram but different word recognition into distinct groups; it changed continuously rather than revealing hidden clusters. Whether these gradients correspond to clinically meaningful subgroups cannot be settled from the published reference schemes and is addressed in separate work.

### Reference schemes name regions of a continuum

Shown together in the same space, the four published schemes look less like competing classifications than like different ways of drawing boundaries on the same territory (Figures 3, 6 and S3). Each has distinct centroids, but the surrounding point clouds overlap without clear gaps, so each names regions of a continuum rather than isolating natural groups, exactly as the continuous descriptions of Allen and Eddins (2010) and Vaden et al. (2022) would predict. Under their native criteria, the schemes left between 11.7% (Parthasarathy) and 36 – 40% (Bisgaard, Dubno, BEAR) of ears unclassified, but these fractions reflect how each scheme was applied more than the phenotypes themselves. The rule for assigning an audiogram to each scheme was our decision, not one prescribed by the original studies, and Parthasarathy’s fuller coverage reflects its more permissive rule as much as clusters learned from a cohort like ours.

Re-classifying all four under a single common rule confirmed this: the schemes with wider native tolerances (Dubno, BEAR and Parthasarathy) lost far more ears, whereas Bisgaard, which already uses that band, was unchanged. Because the common band is narrower than the Dubno and BEAR ranges, it improves comparability at the cost of fidelity. Since no single rule achieves both at once, we kept the native criteria for the figures. This carries a clear implication for any future data-driven method: a clustering approach should provide not only its phenotype means and limits but also a rule for assigning new, unseen ears, derived from and consistent with the same model, so that coverage can be compared fairly across datasets.

The schemes also differ in the data they were built from and in their intended use, which limits how completely any of them can describe an unselected clinical population. Their origins range widely: Gaussian-mixture clustering of 116,400 clinical audiograms (Parthasarathy et al. 2020), vector quantisation of 28,244 audiograms for hearing-aid design (Bisgaard et al. 2010), animal models of age-related cochlear pathology (Dubno et al. 2013), and an extensive test battery including suprathreshold measures in only 75 human listeners (Sanchez-Lopez et al. 2020). Under any criterion, a large fraction of ears falls outside all published categories, which supports a representation of the audiological space that does not depend on a single scheme, in line with efforts to harmonise auditory profiles across clinics (Saak et al. 2022, 2025).

#### Towards a Hearing Loss Map

Together, these results point towards a map rather than a fixed set of phenotypical categories. The audiological UMAP is our first version of a *Hearing Loss Map*: a two-dimensional, clinically interpretable space that, unlike a purely audiometric one, is shaped by suprathreshold performance as well as by the audiogram (Figure 6). With severity increasing from west to east and sloping losses towards the north, every ear has a position relative to all the others, and the map begins to represent the variation that the audiogram, and the categories based on it, leave hidden (Kujawa and Liberman 2009; Grant et al. 2022). We use the word “begins” deliberately: with only word recognition in quiet added, the map gains graded structure, but it does not yet separate hearing losses into distinct, mechanism-specific phenotypes. The dataset and non-linear visualisation framework presented here provide the foundation for this map. Developing it into a tool for following patients over time, and potentially for anticipating an individual patient’s hearing-loss trajectory as projected onto the map, is a goal of ongoing efforts in longitudinal, data-driven auditory phenotyping. Realising it will depend on the curated databases and shared data standards now emerging in audiology (Callejón-Leblic et al. 2024; Vercammen et al. 2026).

### Limitations

This work has several limitations. The data are retrospective, from a single centre, and collected over almost thirty years, so heterogeneity is reduced by preprocessing but not removed (Dillard et al. 2020). Because these are routine clinical records rather than data collected for research, we had no control over which tests were performed or how they were administered. This concerns experimental control, not data quality: the measurements were made by trained audiologists at a specialist tertiary centre and used for real diagnostic and treatment decisions, and the trade-off for this loss of control is scale, a cohort far larger than a controlled prospective study could assemble. As a result, only one suprathreshold measure, WRS_max_-Q, was available at scale; speech-in-noise was recorded too rarely to use, so the audiological space we could build is narrow. A more complete test battery would probably reveal more non-linear structure. The tone-plus-speech subset (213,237 of 251,986 ear-session units) excludes ears without a speech score and may be biased towards patients considered to warrant speech testing. More fundamentally, neither PCA nor UMAP can handle missing values, so any ear with an incomplete set of measurements is removed; this forced the complete-case restriction and makes it difficult to add measures that are only partially collected in clinical practice, such as speech-in-noise, acoustic reflex thresholds or physiological responses. Encoders that tolerate missing data, such as variational autoencoders, could use these incomplete measurements and are a natural next step. UMAP is a visualisation tool rather than a true metric: it does not preserve global distances, it depends on its parameters, and its orientation was fixed by us, so it should be read qualitatively and treated as exploratory (McInnes et al. 2020). Finally, the published reference schemes were applied outside the populations in which they were developed. BEAR, originally based on a suprathreshold battery, could only be approximated here from the audiogram. Without histopathological ground truth, our diagnoses rest on the ABG and on ICD-10 codes, which are coarse and mostly non-specific.

### Conclusion

In summary, the pure-tone audiogram of a large clinical population is low-dimensional and continuous, and a linear embedding represents it well. A non-linear embedding becomes necessary only when the data become more audiological in nature, meaning they include suprathreshold information, such as word recognition, that the audiogram cannot predict. Treating auditory phenotyping as a position in a continuous, interpretable space rather than as a member of a discrete category may provide a more faithful representation of a fundamentally graded condition, and offers a possible foundation for the development of a clinically useful Hearing Loss Map.

## Author contributions

**Gerard Encina-Llamas:** Conceptualization; Methodology; Software; Formal analysis; Data curation; Visualization; Writing — original draft, review & editing; Project administration; Funding acquisition. **Erik Kjærbøl:** Resources; Data curation; Writing — review & editing. **Abigail Anne Kressner:** Conceptualization; Methodology; Writing — review & editing; Project administration.

## Statements and Declarations

### Ethical considerations

This retrospective study analysed existing clinical audiological records. As a non-interventional study of existing clinical data, it did not require approval from a research ethics committee; it was authorised by the Center for Regional Udvikling (Capital Region of Denmark) under Pactius project P-2022-629 and conducted in accordance with the Declaration of Helsinki and the GDPR (see *Approvals and data governance*).

### Consent to participate

Individual informed consent was not required for this retrospective analysis of existing clinical records, authorised under Pactius project P-2022-629 (Center for Regional Udvikling, Capital Region of Denmark).

### Consent for publication

Not applicable; no data from an identifiable individual person are presented.

### Declaration of conflicting interests

The authors declared no potential conflicts of interest with respect to the research, authorship, and/or publication of this article.

### Funding

The study was funded by the GN Foundation (Denmark), grant number 273.

### Data availability

The clinical audiological records analysed here are personal health data and cannot be made publicly available: under the governing data-access agreement and applicable data-protection regulation (GDPR), they were processed only within the secure DTU Computerome HPC environment and cannot be shared or exported. The analysis code is not publicly released but is available from the corresponding author on reasonable request for non-commercial research.

## Supplementary Material

**Figure S1.**
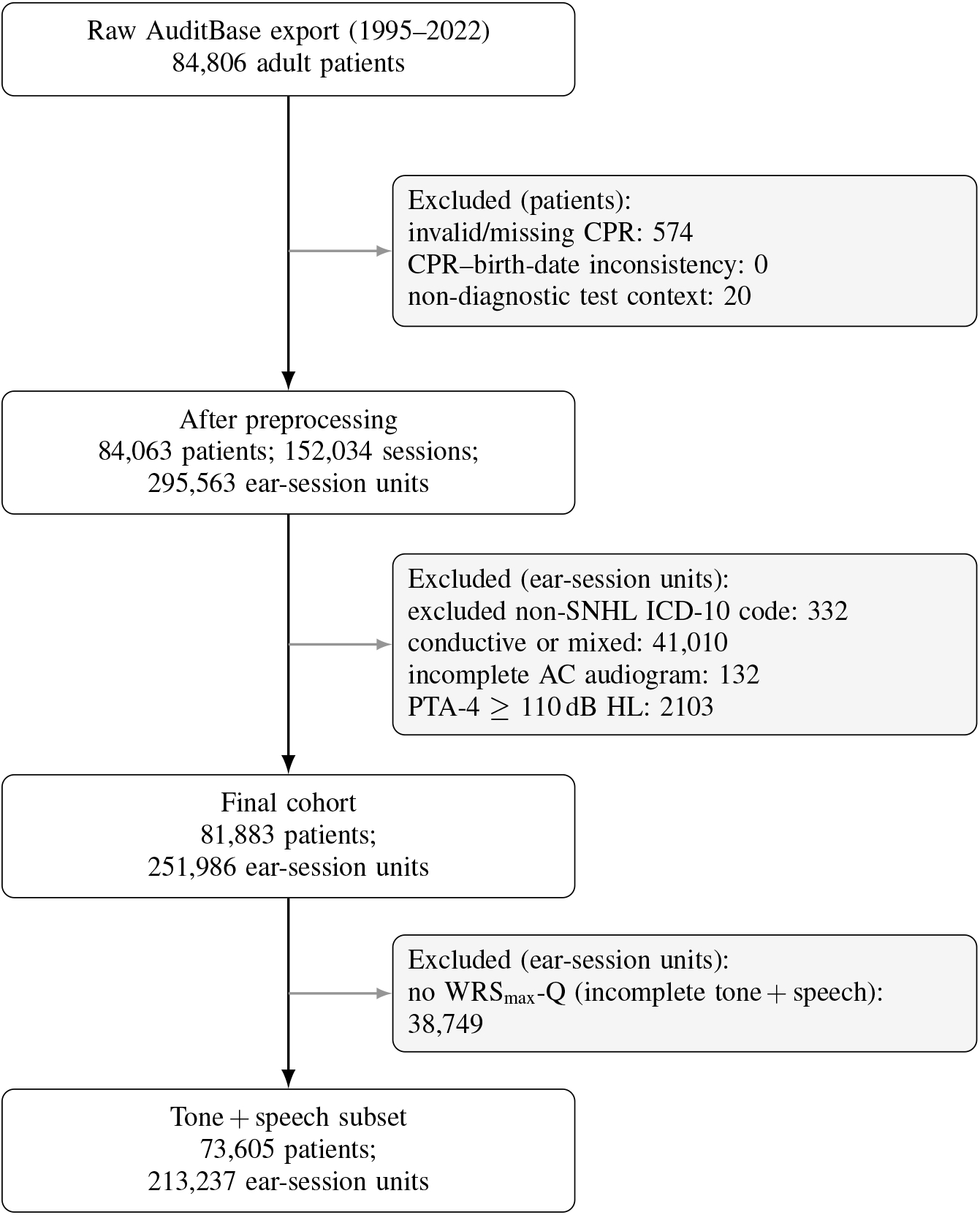
Cohort attrition diagram. Left column: numbers retained after each preprocessing and final-cohort step (patients, sessions and ear-session units). Grey boxes: per-step exclusions, counted as patients for the preprocessing step and as ear-session units for the final-cohort and tone + speech steps.

**Table S1.** Distribution of ear-session units across published reference schemes in the final cohort (251,986 ear-session units). Values are counts and percentages.

| Reference scheme | Category | *n* (ear-session units) | % |
| --- | --- | --- | --- |
| Bisgaard et al. (2010) | N0 | 23,793 | 9.4 |
|  | N1 | 25,441 | 10.1 |
|  | N2 | 27,207 | 10.8 |
|  | N3 | 37,300 | 14.8 |
|  | N4 | 14,873 | 5.9 |
|  | N5 | 3764 | 1.5 |
|  | N6 | 1393 | 0.6 |
|  | N7 | 889 | 0.4 |
|  | S1 | 9227 | 3.7 |
|  | S2 | 4725 | 1.9 |
|  | S3 | 5709 | 2.3 |
|  | Unclassified | 97,665 | 38.8 |
| Dubno et al. (2013) | Old Normal Hearing | 18,605 | 7.4 |
|  | Pre-metabolic | 17,032 | 6.8 |
|  | Metabolic | 36,608 | 14.5 |
|  | Sensory | 16,282 | 6.5 |
|  | Metabolic-Sensory | 69,539 | 27.6 |
|  | Unclassified | 93,920 | 37.3 |
| BEAR (Sánchez-López et al., 2020) | Profile A | 64,332 | 25.5 |
|  | Profile B | 25,295 | 10.0 |
|  | Profile C | 46,517 | 18.5 |
|  | Profile D | 15,443 | 6.1 |
|  | Unclassified | 100,399 | 39.8 |
| Parthasarathy et al. (2020) | M1 | 9954 | 4.0 |
|  | M2 | 3775 | 1.5 |
|  | M3 | 28,638 | 11.4 |
|  | M4 | 3473 | 1.4 |
|  | M5 | 7024 | 2.8 |
|  | M6 | 27,448 | 10.9 |
|  | M7 | 65,552 | 26.0 |
|  | M8 | 4793 | 1.9 |
|  | M9 | 23,551 | 9.3 |
|  | M10 | 48,295 | 19.2 |
|  | Unclassified | 29,483 | 11.7 |

**Figure S2.**
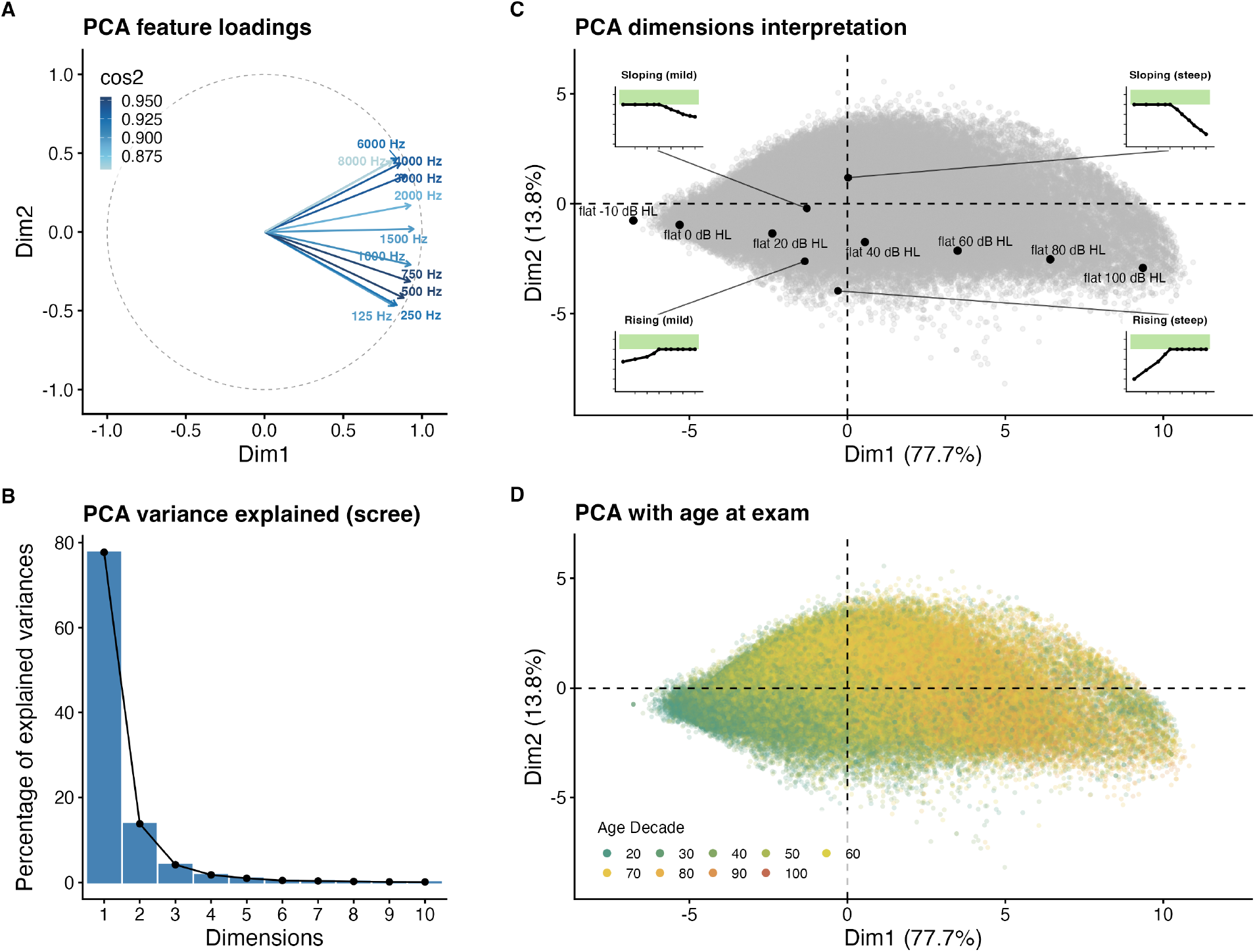
Interpretation of the PCA of the final cohort. (A) Feature loadings (colour: cos^2^): all eleven frequencies load positively on Dimension 1, while high frequencies load positively and low frequencies negatively on Dimension 2. (B) Scree plot of the variance explained, dominated by the first two components. (C) Synthetic audiograms projected into the space: flat audiograms of increasing level (−10 to 100 dB HL) move along Dimension 1, whereas sloping (high-frequency loss) and rising (low-frequency loss) audiograms are displaced towards positive and negative Dimension 2, respectively (insets show the audiogram shapes). (D) Individual ears coloured by age decade at examination.

**Figure S3.**
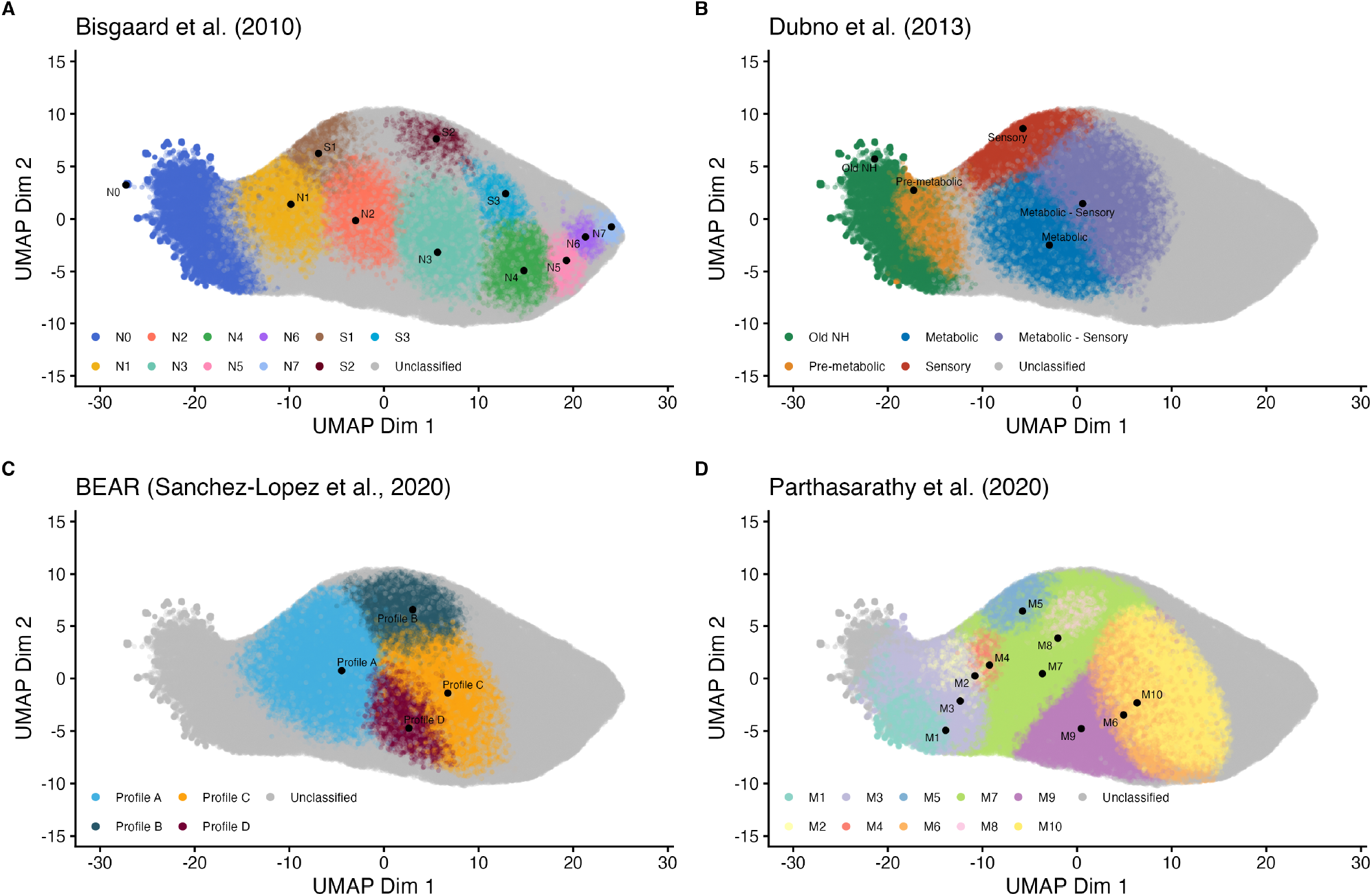
UMAP embedding of the final cohort using the eleven AC thresholds (tone-only feature set). Panels and colouring mirror Figure 3: (A) Bisgaard; (B) Dubno; (C) BEAR; (D) Parthasarathy. Dimension 1 is signed to increase with PTA-4; Dimension 2 with the HF–LF slope.

**Figure S4.**
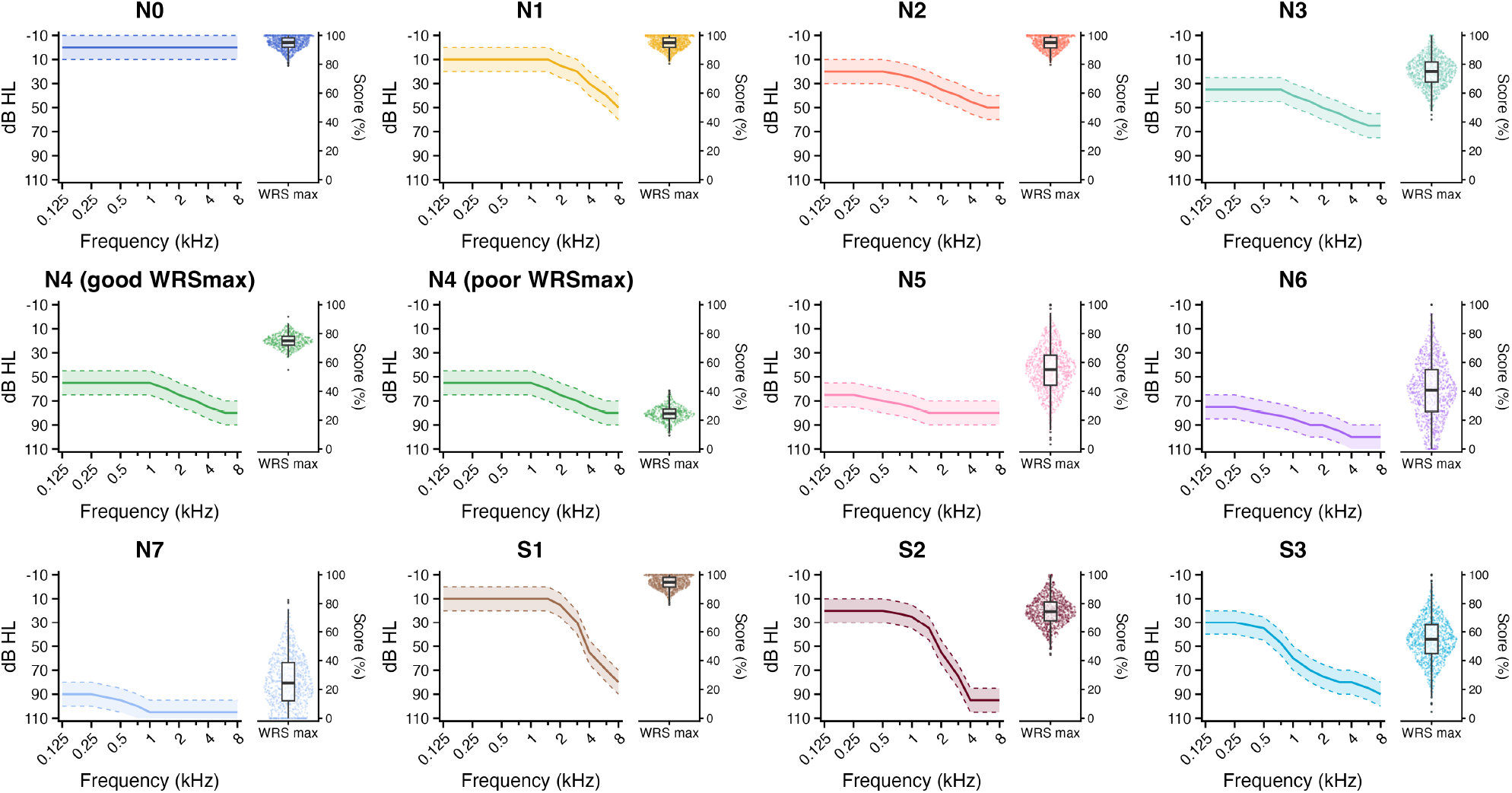
Synthetic reference data for the demonstration in Figure 5. For each Bisgaard profile (N0–N7, S1–S3), synthetic AC audiograms were drawn from the profile mean *±* range (left) and a maximum word-recognition score (WRS_max_) assigned from a realistic, profile-specific distribution (right, violin with boxplot). Profile N4 is split into two subgroups with identical audiograms but good versus poor WRS_max_.

## Notes

### Competing Interest Statement

The authors have declared no competing interest.

### Author Declarations

This retrospective study analysed existing clinical audiological records. As a non-interventional study of existing clinical data, it did not require approval from a research ethics committee; it was authorised by the Center for Regional Udvikling (Capital Region of Denmark) under Pactius project P-2022-629 and conducted in accordance with the Declaration of Helsinki and the GDPR.

